# Simulate Scientific Reasoning with Multiple Large Language Models: An Application to Alzheimer’s Disease Combinatorial Therapy

**DOI:** 10.1101/2024.12.10.24318800

**Authors:** Qidi Xu, Xiaozhong Liu, Xiaoqian Jiang, Yejin Kim

## Abstract

**Motivation:** This study aims to develop an AI-driven framework that leverages large language models (LLMs) to simulate scientific reasoning and peer review to predict efficacious combinatorial therapy when data-driven prediction is infeasible.

**Results:** Our proposed framework achieved a significantly higher accuracy (0.74) than traditional knowledge-based prediction (0.52). An ablation study highlighted the importance of high quality few-shot examples, external knowledge integration, self-consistency, and review within the framework. The external validation with private experimental data yielded an accuracy of 0.82, further confirming the framework’s ability to generate high-quality hypotheses in biological inference tasks. Our framework offers an automated knowledge-driven hypothesis generation approach when data-driven prediction is not a viable option.

**Availability and implementation:** Our source code and data are available at https://github.com/QidiXu96/Coated-LLM

## Introduction

Recent advancements in large language models (LLMs) have demonstrated their disruptive potential in scientific discovery. These models have efficiently tackled combinatorial optimization problems, often surpassing traditional heuristics [1,2]. LLMs have also shown success in various chemistry and materials science tasks, such as predicting molecular properties and chemical reaction yields [3], and autonomously searching for chemicals [4,5]. Our research aligns with efforts to develop “autonomous scientists” but with a unique focus on mimicking human’s scientific reasoning.

In scientific investigation, researchers ask questions to understand the underlying mechanisms of phenomena, applying deductive reasoning to derive specific predictions from general principles and inductive reasoning to form general conclusions based on specific observations [6]. Hypotheses generated through this reasoning are tested experimentally to confirm or refute them. However, traditional scientific reasoning is often limited by human bias and cognitive capacity [6]. Researchers may exhibit confirmation bias, favoring data that supports their preconceptions, and struggle to process the vast amount of existing literature, leading to incomplete reviews and overlooked insights. Additionally, managing multiple factors and identifying subtle patterns often exceeds human cognitive abilities.

An automated approach using artificial intelligence (AI), particularly LLMs, can mitigate these limitations by assisting human scientific reasoning, as evidenced in several prior studies [7–9]. This study systematically explores the potential of LLMs to automate hypothesis generation (i.e. prediction) via scientific reasoning. While data-driven machine learning models can be effective for this purpose when abundant data is available, our focus is on scenarios where such data is scarce, which is more common in the real world. We chose a specific application that necessitates a deep understanding of domain knowledge and capability for critical reasoning beyond mere memorization: identifying effective combinations of drugs for in vivo experiments in complex systemic diseases. We chose to focus on Alzheimer’s disease (AD), a complex neurodegenerative condition where multiple disease etiologies entangle together, thus a comprehensive consideration of multiple underlying mechanisms is critical when developing therapeutics.

Drug combination therapy is to use of two or more therapeutic agents to treat a single disease. This approach is particularly prevalent in the treatment of complex diseases such as diabetes and metabolic syndrome, cardiovascular disease, cancer, and others [10–17]. The goal of combination therapy is to achieve a more effective treatment outcome than what could be achieved with a single drug. Developing effective combinatorial therapies faces significant challenges, particularly in selecting the right therapeutic agents (drugs) and relevant in vivo models. Researchers make specific predictions about the potential efficacy of various combinations from general principles and known mechanisms of action (deductive reasoning). The complexity arises from the factorial growth in the number of possible combinations (therapeutic agent 1 * therapeutic agent 2 * in vivo model), making it impractical for human experts to manually evaluate all possibilities. Also, researchers form general conclusions about the effectiveness of these combinations based on observations of similar combinations. This inductive reasoning is particularly challenging due to the variability of therapeutic agents and in vivo models. For example, while some in vivo models are well-established (e.g., 3xTg-AD), many others are developed ad hoc (e.g., AlCl₃-fed rats, Hyperhomocysteinemia (HHcy)-induced AD rat model). Similarly, new therapeutic agents are continuously being developed, making it difficult for researchers to derive patterns to draw conclusions.

Only a few studies have explored the use of LLMs for combinatorial search problems. A recent study [9] introduced CancerGPT, an LLM-based model designed for predicting drug pair synergy in rare tissues with limited data. It demonstrates the capability of LLMs to handle complex biological inference tasks. However, CancerGPT primarily focuses on finetuning LLMs using high-throughput in vitro experimental data. Such data is not available for most complex diseases in which no cell line models can represent the multiple systems that complex disease affects simultaneously. In contrast, our proposed framework, **Co**mbinatorial **A**lzheimer’s disease **T**herapeutic **E**fficacy **D**ecision (Coated-LLM), avoids such resource-intensive finetuning by leveraging systematic in-context learning.

In addition, combination therapy poses greater challenges for an LLM alone because the multiway interactions between therapeutics and in vivo models add layers of complexity to the reasoning process. By integrating multiple LLM agents simulating scientific peer review (*Researcher*, *Reviewer*, *Moderator*) and injecting external knowledge, our approach aims to mimic human scientific reasoning, which is more flexible and versatile than task-specific machine learning models. Comprehensive computational evaluation demonstrates that our framework identifies potent combinations, thus assisting human scientists to scale up scientific reasoning.

## Results

### Model summary

We create an AI model *f* that automates scientific reasoning to generate hypotheses on efficacious combinatorial therapy for in vivo experiments (Algorithm 1, Fig 1). Mimicking the scientific discovery process of human researchers, our framework consists of multiple LLM agents playing different roles: *Researcher*, *Reviewer*, and *Moderator*. *Researcher* generates a series of reasoning steps to propose a prediction on the efficacy of combinatorial therapeutic agents. Multiple *Reviewers* review and criticize the quality of the prediction generated by the *Researcher* and offer feedback. Finally, *Moderator* integrates the *Researcher’s* proposed prediction and the *Reviewers’* feedback to suggest a more valid prediction.

**Figure 1.**
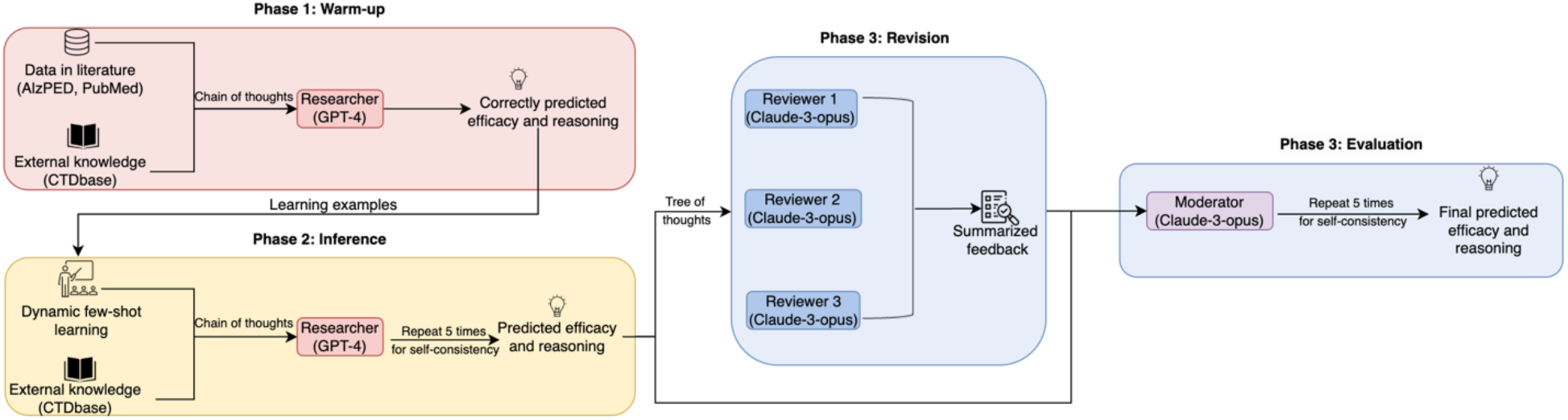
Study overview. Coated-LLM is a structured framework that mimics human scientific reasoning and peer review processes to generate hypotheses on efficacious combinatorial therapy. It consists of three stages: (i) Warm-up phase, where *Researcher* uses external biological knowledge to practice scientific inference and keep correct predictions as learning examples. (ii) Inference phase, where *Researcher* inferences the new combination using its top five similar questions from learning examples and gets the consistency prediction. (iii) Revision phase, where multiple *Reviewers* provide feedback and *Moderator* integrates consistency prediction from *Researcher* and feedback from *Reviewer* to generate the final consensus prediction.

Throughout all the communication among LLM agents, we prompt the LLM agents to utilize various in-context learning techniques such as integrating external biomedical knowledge, few-shot learning [18], self-generated chain-of-thoughts (CoT) [19] and/or tree of thoughts (ToT) [20], and self-consistency [21]. See method details at Warmup, Inference, and Revision phases.

### Data collection and augmentation

Historical data on effective drug combinations for AD are very scarce. For the purpose of evaluation and providing a few learning examples, we collect the data via literature mining (see Supplementary B). This is to leverage in-context learning rather than finetuning an LLM or building a supervised model. As a result, we identified a total of 242 articles reporting 250 drug combinations with positive efficacy and 30 with negative efficacy (Fig. 2a). Fig 2b summarizes the top five most frequently mentioned terms across therapeutic agents, animal models, and pathways.

**Figure 2.**
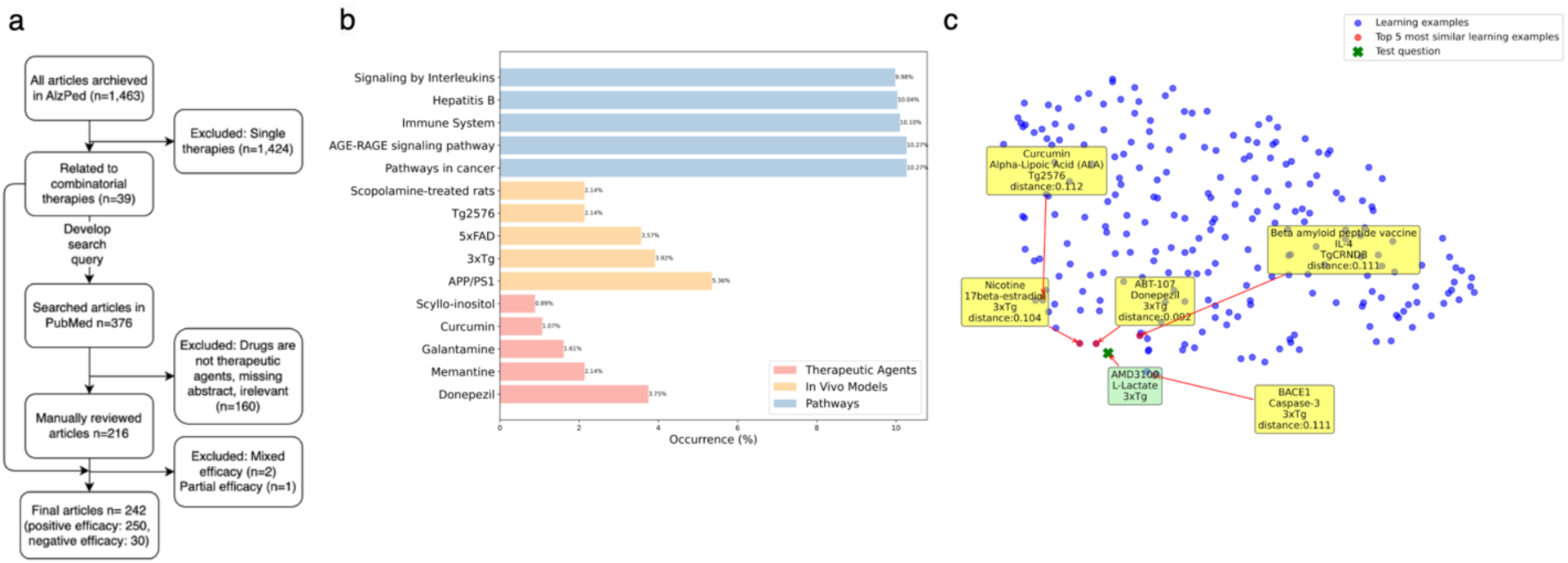
Distribution of drug combinations and efficacy in literature. **a**. Data collection from literature. The process began with an initial pool of articles from the AlzPED, followed by additional searches conducted in PubMed. Articles were screened and excluded based on predefined criteria. The final selected literatures included articles that reported drug combinations with positive or negative efficacy. **b**. Top 5 frequent terms in therapeutic agents, animal models, and pathways. **c**. UMAP visualization of drug combinations and efficacy. Each drug combination is converted into a natural language question to generate embeddings with OpenAI’s text-embedding-ada-002 model. The UMAP projection, derived from these embeddings, reveals that the combination (AMD3100, L-Lactate, 3xTg), for example, is similar to combinations which have same animal model (e.g., ABT-107, Donepezil, 3xTg).

Given the fact that only a small portion of drug combinations show positive efficacy out of the majority of drug combinations by nature, our collection of prior literature has a reporting bias toward positive efficacy (Fig. 2a). Prior researchers were more incentivized to report positive outcomes, rather than negative outcomes. However, it is equally important for our model to detect combinations that may not have positive efficacy. Utilizing this biased data in our model development would result in biased prediction. Therefore, we augmented the data with unlabeled combinations and regarded the unlabeled data as noisy data with non-positive efficacy (see Supplementary C). After data augmentation, we have a total of 530 combinations (250 combinations with positive efficacy; 30 combinations with negative efficacy, augmented 250 combinations with noisy, non-positive efficacy).

We then extracted pathway information for drugs after comparing different modalities (such as gene names, gene interaction information, and phenotypes) as a retrieval augmented generation (RAG) [22], see Retrieval augmented generation section. Of the total combinations, 129 had pathway information for both drugs available from CTDbase [23], and 235 had information for only one of the two drugs.

### Coated-LLM achieved significant accuracy in cross validation

We developed and evaluated Coated-LLM using the data collected and augmented from literature. In the Warmup phase, we first created learning examples for the *Researcher* to utilize during few-shot learning of the next phase. We set aside 70% of the whole data, kept combinations that the *Researcher* can predict correctly, and used them as learning examples. Finally, 231 combinations were predicted correctly (134 combinations with positive efficacy; 97 combinations with non-positive efficacy). We converted these combinations (i.e., Drug 1, Drug 2, animal model) into natural text questions (see Supplementary A, example at Supplementary F). The questions, generated CoT reasonings (See Warm-Up phase, Chain-of-thoughts section), and efficacy answers (positive or non-positive) were subsequently incorporated as learning examples for the Inference phase (see Warm-Up phase section, example at Supplementary G).

In the Inference phase, we first identified most relevant examples for each combination of interest for few-shot learning. Each combination was converted into a corresponding question, similar to the Warm-Up phase. For each question, we identified the top five most similar questions from the learning examples. These selected questions, along with their corresponding generated CoT reasonings and efficacy answers, were then utilized for few-shot learning (see Inference phase, Dynamic Few-shot section). To demonstrate that the selected questions were more relevant to the test questions than other questions from the learning examples, we calculated the mean cosine distance between the test question embeddings and learning examples embeddings. The mean top five average cosine distance was 0.08 (variance: 0.0002), while the mean overall average cosine distance was 0.13 (variance: 0.0003). The results indicate that test questions are significantly relevant to specific questions in the learning examples than to the overall set of learning examples. Figure 2c provided a visual representation of similarities between the target combination and learning examples. Later, we replicated the few-shot learning process five times for each question to achieve a majority-vote efficacy answer and the most detailed generated CoT reasoning (see Inference phase, Self-consistency via ensemble section).

In the Revision phase, previous phase generated reasonings were subsequently forwarded to the *Reviewer* for comprehensive feedback (see Revision phase, Evaluate section, example at Supplementary H). To obtain the final consensus efficacy answers for each combination of interest, we integrated the most detailed CoT reasonings from the Inference phase with feedback from the *Reviewer*. This compiled data was processed through the *Moderator* LLM five times to get the final consensus efficacy answers (see Revision phase, Revise section, example at Supplementary I). Among the total 156 combinations of interest, 129 demonstrated more than 80% consistency across 5 rounds, with 83 of them achieving 100% consistency.

The final prediction achieved an accuracy of 0.74, with a precision of 0.71, a recall of 0.79, and an F1-score of 0.75. Table 1 presents the contingency table for Coated-LLM’s predictions and examples of misclassifications. These results indicate that our model is effective in predicting the efficacy of drug combinations for Alzheimer’s Disease, exhibiting a balanced performance with both high precision and recall. Compared to the traditional experimental screening approach, Coated-LLM enhances scientific reasoning capabilities and introduces a novel, time-efficient, and cost-effective approach for identifying potential therapeutic agent combinations.

**Table 1.** Contingency table of prediction outcomes with examples using Coated-LLM.

| **Drug A** | **Drug B** | **Animal model** | **Predicted efficacy** | **Actual efficacy (reference)** |
|---|---|---|---|---|
| **True positive (n=61)** |  |  |  |  |
| Lycopene | Vitamin E | Tau P301L | Positive | Positive [27] |
| Donepezil | Fluoroethylnormemantine | 5xFAD | Positive | Positive [28] |
| **False positive (n=25)** |  |  |  |  |
| Cholesterol | Homocysteine | Sprague Dawley rats | Positive | Non-positive [29] |
| PNU-120596 | Fluoxetine | Scopolamine-treated Rats | Positive | Non-positive (Augmented data) |
| **False negative (n=16)** |  |  |  |  |
| Gallic Acid | Sodium Arsenite | Male rats | Non-positive | Positive [30] |
| (m)RVD-hemopressin | AM251 | SH-SY5Y cells | Non-positive | Positive [31] |
| **True negative (n=54)** |  |  |  |  |
| Atorvastatin | Farnesol | C57BL/6 | Non-positive | Non-positive [32] |
| Galantamine | Mecamylamine | ICR | Non-positive | Non-positive [33] |

In comparison, the performance metrics for the baseline model (network-based approach, see Baselines) achieved an accuracy of 0.52, precision of 0.46, recall of 0.16, and an F1 score of 0.24 (Table S1). The baseline model exhibits significant limitations, particularly it does not accommodate therapeutic agents that are not conventional drugs, such as membrane-free stem cell extracts, for which gene target data are typically unavailable.

Interestingly, the augmented (non-positive) data that were predicted to be positive may suggest intriguing hypotheses. Specifically, the Coated-LLM generated 25 instances that were false positives. These instances included combinations that were predicted to have positive efficacy despite being labeled as non-positive in our augmented dataset. Although these combinations were marked as false positives, we hypothesize that they may, in fact, be efficacious combinations that have not yet been experimentally tested. One such combination is PNU-120596 and Fluoxetine in Scopolamine-treated Rats. This combination leverages the cognitive-enhancing effects of PNU-120596, a positive allosteric modulator of α7-nAChR [24], with Fluoxetine, an SSRI known for its role in neurogenesis and synaptic plasticity [25]. The combination could theoretically improve memory function by potentially enhancing cholinergic and serotonergic neurotransmission. Moreover, Fluoxetine’s inhibition of the cytochrome P450 enzyme CYP2D6 [26] may result in elevated plasma levels of PNU-120596, potentially amplifying its cognitive-enhancing effects. To further explore these possibilities, we are preparing in vivo experiments to evaluate the efficacy of these false positive combinations. These studies will provide crucial data to verify our model’s predictions.

### Ablation Study

Aiming to understand the contributions of each component within our model, we conducted an ablation study (Fig. 3). The ablation study begins with zero-shot learning, where GPT-4 leverages its pre-learned knowledge to make predictions. Introducing dynamic few-shots [1] results in a slight performance decrease, likely due to probable mislabeled augmented combinations used as few-shot examples. Subsequently, applying RAG, we integrated external biomedical knowledge on pathway, which led to significant improvements in accuracy (+17%), precision (+13%), recall (+40%), and F1-score (+23%). By implementing self-consistency via an ensemble strategy, we achieved consistent and reliable predictions and further increased the accuracy by 6%, precision by 4%, recall by 3%, and F1-score by 4%. Finally, incorporating feedback from other LLM agents such as *Reviewer* and *Moderator* further improved the model’s performance, correcting potential errors to achieve the highest accuracy (+5%) and precision (+9%). However, we observed a decrease in recall due to *Reviewer* and *Moderator*, suggesting that the two LLM agents in Revision phase favor reducing false positive (the review process is more skeptical and stringent). Despite the decrease in the recall, we kept the *Reviewer* and *Moderator* because, in real in vivo experiments, our model is used to retrieve a ranked list of probable positive combinations, thus high precision is more important than high recall.

**Figure 3.**
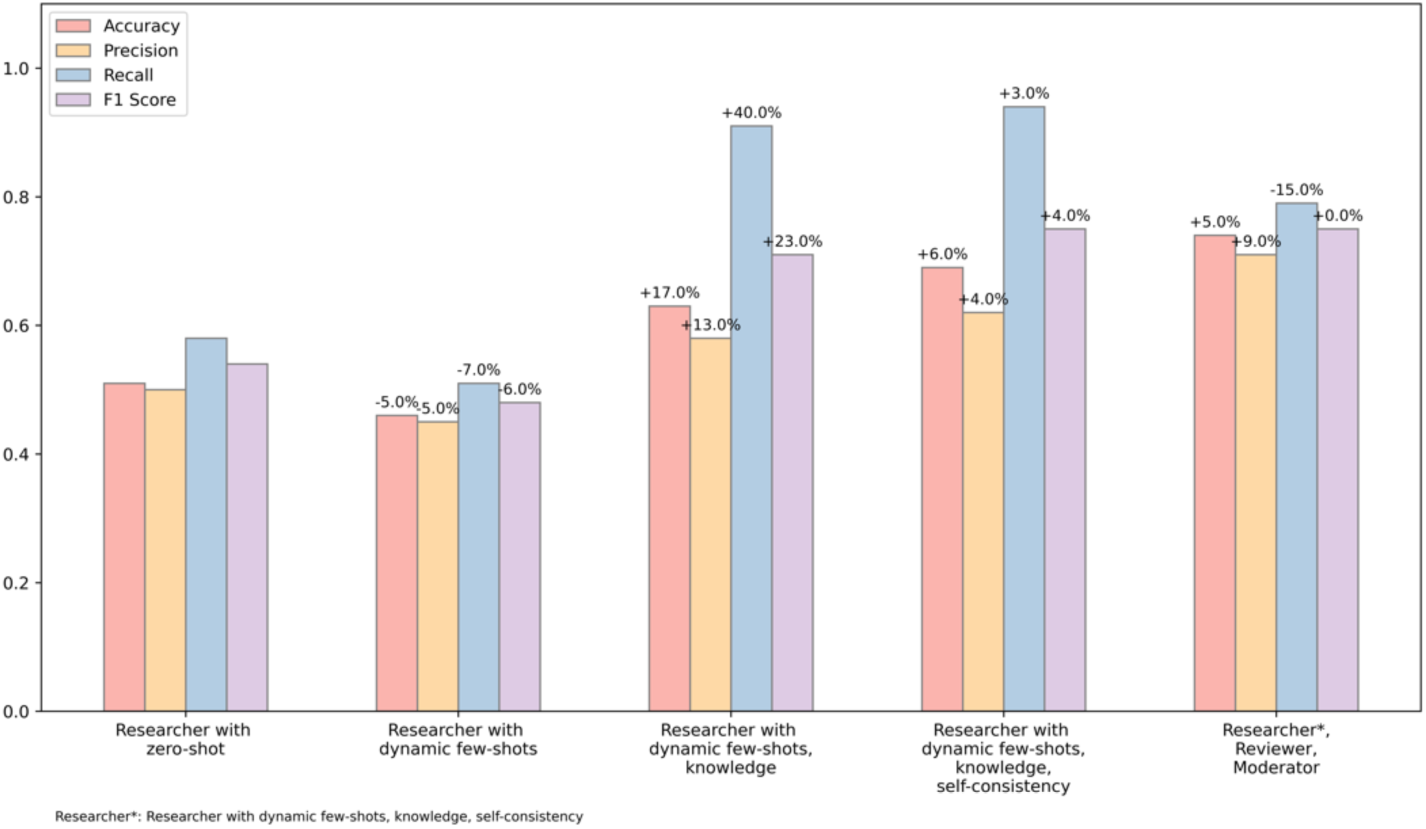
Visual illustration of Coated-LLM components and additive contributions to the performance. Coated-LLM combines kNN-based five-shots dynamic learning example selection, external pathway knowledge, self-consistency (n=5), *Reviewer*, and *Moderator*.

### Coated-LLM achieved significant accuracy in external validation

We also utilized our private dataset to evaluate our model’s capabilities. This evaluation is crucial as the external data remains private, eliminating any risk of data leakage through GPT-4. Additionally, the dataset is independent of the training and testing datasets, ensuring no overlap. The external data set included nine non-positive and two positive drug and cell line combinations. Unlike the 1:1 ratio of positive to non-positive samples in the cross-validation test set, the external validation set had a more challenging and realistic ratio of 1:4.5, with nine non-positive samples out of eleven (Supplementary D). This higher ratio reflects the rarity of efficacious and synergistic drug combinations in the real world, making accurate predictions more difficult compared to the cross-validation set.

As a result, due to the distribution shift between (cross-validation) test set and the external set, the prediction task was indeed challenging. Our model demonstrated an accuracy of 0.82, with a precision of 0.50, a recall of 0.50, and an F1-score of 0.50 (Table 2). In comparison, the performance metrics for the baseline model (Table S2) achieved an accuracy of 0.27.

**Table 2.** Contingency table of Coated-LLM’s prediction for the external data with examples.

| **Drug A** | **Drug B** | **Model** | **Predicted efficacy** | **Actual efficacy** |
|---|---|---|---|---|
| **True positive (n=1)** |  |  |  |  |
| Galantamine | Caffeine | HT22 Mouse Hippocampal Neuronal Cell Line | Positive | Positive |
| **False positive (n=1)** |  |  |  |  |
| Donepezil | Salicylic | HT22 Mouse Hippocampal Neuronal Cell Line | Positive | Non-positive |
| **False negative (n=1)** |  |  |  |  |
| Galantamine | Mifepristone | HT22 Mouse Hippocampal Neuronal Cell Line | Non-positive | Positive |
| **True negative (n=8)** |  |  |  |  |
| Galantamine | Diclofenac | HT22 Mouse Hippocampal Neuronal Cell Line | Non-positive | Non-positive |
| Rivastigmine | Lithium | HT22 Mouse Hippocampal Neuronal Cell Line | Non-positive | Non-positive |

## Discussion

In this study, our framework leverages the strengths of prompt engineering to mimic human scientific reasoning processes. By incorporating multiple agents such as *Researcher*, *Reviewer*, and *Moderator*, our approach not only scales up the scientific reasonings but also realizes the hypotheses on efficacious combinatorial therapy in an automated way. The Coated-LLM achieved notable accuracy in both held-out test set and external set indicating its effectiveness in predicting the efficacy of drug combinations in the wild. The following lessons were derived from our study:

- Interaction of Multi-Agent LLM: Coated-LLM introduces a tripartite agent system wherein the Researcher LLM employs structured prompt-driven CoT reasoning to generate hypotheses. The Reviewer LLM, utilizing the ToT strategy, provides essential feedback, enhancing the completeness of the generated hypotheses. Meanwhile, the Moderator LLM plays a crucial role in refining these hypotheses in a skeptical and conservative way, ensuring reliability of hypotheses. The collective integration of multiple LLM agents introduces a robust system that enhances the quality and accuracy of hypotheses generation.
- Implications of Dynamic Few-Shot Learning: Dynamic few-shot learning plays a crucial role in enhancing the predictive capabilities of LLM by leveraging examples that are most similar to the target data. Our findings reveal that the inclusion of high-quality real learning examples significantly enhances the accuracy of predictions compared to zero-shot strategy. In contrast, the use of augmented learning examples does not yield a similar increase in accuracy, showing an intriguing observation that real data examples are more beneficial than the augmented data for improving model performance in dynamic few-shot learning contexts. In addition, our study further underscores the importance of maintaining a balanced set of learning examples. A predominance of examples from a singular class within the dynamic few-shot learning examples may make biased predictions, highlighting the need for balance in learning examples.
- Enhancements Through Retrieval-Augmented Generation (RAG): The adoption of RAG, which integrates domain-specific knowledge is crucial. This strategic inclusion bridges the inherent knowledge gaps within the pre-trained GPT-4, resulting in significant enhancements in precision, recall, accuracy, and F1-score.
- Stabilization via Ensemble Methods: Employing ensemble methods mitigates the risk associated with a single prediction, providing a more stable and consistent output.

It is important to acknowledge that the external validation is limited by the small and private dataset. However, the inclusion of the cross-validation test set provides a complementary evaluation, supporting the generalizability of our findings. Given that the initial learning examples contain only 97 non-positive efficacy combinations out of 231 (41.9%), such bias presents a considerable challenge for our model in accurately predicting non-positive outcomes. Despite these challenges, our model achieved a significant accuracy rate, demonstrating its effectiveness in generating hypotheses for synergistic drug combinations with minimal historical data.

While we have obtained promising results, our study has several limitations. One of the primary limitations is the underrepresentation of negative combinations within the dataset obtained through literature mining. This disproportion led to a predominance of positive combinations in the learning examples, subsequently introducing a bias during the model’s Inference and Revision phase. Such a bias tends to skew predictions toward more frequently observed positive outcomes, potentially compromising the model’s accuracy in identifying truly efficacy combinations. To mitigate this issue, we implemented a strategy of data augmentation. While this approach helped balance the dataset and was designed to better reflect the real-world scenario, where negative combinations are more prevalent than positive ones, it is important to acknowledge that these augmented combinations could be false negatives. This mislabeling could lead to incorrect decisions about the efficacy of drug combinations. For comparison, we conducted the ablation study without negative data augmentation (Fig S1). In the absence of augmentation, we observed a clear accuracy improvement thanks to the dynamic few-shot strategy. However, the models were overly optimistic and predicted results were biased toward positive efficacy.

Another limitation is that Coated-LLM relies on existing knowledge of drugs. Many combination therapies may arise from unknown activities, such as off-target effects or complex interactions between drug metabolites, which are not captured by current datasets or domain knowledge. Therefore, our approach is limited in identifying combinations that drugs are uncovered or less explored.

While Coated-LLM has been effective in identifying the efficacy of drug combinations for Alzheimer’s disease, its applicability extends to other diseases as well. Exploring its use in other complex diseases could further enhance the application of the Coated-LLM. Additionally, incorporating feedback from real-world applications and clinical trials may provide valuable insights and guide further improvements.

## Methods

### Warm-Up phase

#### Overview

In the warm-up phase, *Researcher* generates answers to training questions and compares them with ground truth answers. The correctly generated answers are used as learning examples in the next inference phase. For this purpose, we split the data into 70% training and 30% test sets, which are used to derive learning examples in the warm-up phase and actual inference in the next phase, respectively. This training set is not for actual training nor fine-tuning the LLM but for learning examples. We set aside a higher proportion for training to ensure the *Researcher* can be exposed to diverse learning examples.

#### Chain-of-thoughts (CoT)

To improve the reasoning ability of the *Researcher*, we applied a chain-of-thought (CoT) prompting strategy by incorporating the instruction: “Take a deep breath and work on this problem step-by-step.” [34]. This approach was designed to encourage the *Researcher* to decompose complex tasks related to the efficacy of drug combinations into a series of intermediate steps, such as identifying drug targets and mechanisms of action, analyzing biological pathways, evaluating multi-pathway targeting, before reaching a final conclusion. (Supplementary F).

#### Retrieval augmented generation (RAG)

To make the *Researcher* answers a question *q* in the training set more intelligently, *Researcher* is allowed to use external biomedical knowledge. Information on therapeutic agents is vast and continuously expanding. LLM generates responses based on patterns learned during training, which are inherently limited by the data they were trained on. The retrieval-augmented generation (RAG) complements the static parameters in LLMs with up-to-date and dynamic information [22]. We retrieve and provide specific external information *Bq* on therapeutic agents *t_1_*, *t_2_*. We used the Comparative Toxicogenomics Database (CTDbase) [23], a knowledge database encompassing 88,144,004 relationships in chemicals, genes, pathways, and diseases. We particularly focused on the pathway information that the therapeutic agents target. Only pathways with a corrected p-value below 0.01 are incorporated as external knowledge, emphasizing their significant enrichment among the genes interacting with the drug. For example, for the therapeutic agent Galantamine, we provided molecular pathway information such as “*Galantamine has several pathway information, such as cholinergic synapse, transmission across chemical synapses, highly calcium permeable postsynaptic nicotinic acetylcholine receptors, …, and peptide hormone metabolism.*” Note that we also tried to incorporate a list of targeting genes as external knowledge, and it did not provide high-quality answers due to its high sparsity. Also note that the in vivo model information, such as one available in AlzForum [35], marginally increased the generation quality while consuming many tokens.

Based on the targeting pathway information *Bq*, we prompt *Researcher* LLM to generate a hypothesis. This hypothesis consists of a series of CoT reasoning (*Cq*) and a final binary answer *Aq* (Supplementary F). If the answer *Aq* is correct and the reasoning *Cq* is logical, we keep the question *q*, reasoning *Cq*, and answer *Aq* in memory to serve as learning examples for the next inference phase. We only focused on the correct answers and filtered out *Cq* if the answer *Aq* is different from the ground truth efficacy label *y*. This simple filtering has greatly decreased the low-quality chain-of-throughs examples [1]. We used GPT-4 for *Researcher* LLM. To encourage *Researcher* LLM to be skeptical, we added the statement ’It is rare for combinations of two drugs to be efficacious and synergistic in real world’ into the prompt (Supplementary F).

### Inference phase

#### Overview

Using the learning examples from the warm-up phase, *Researcher* generates hypotheses to the input questions in the test set. In the inference phase, *Researcher* leverages the learning examples (dynamic few-shot learning) in addition to external biomedical knowledge (RAG), following the same methodology as in the warm-up phase.

#### Dynamic Few-shot

When human researchers are asked a scientific question, human researchers look for similar questions that were answered previously and perform inductive reasoning. So does *Researcher* by leveraging dynamic few-shot learning. Few-shot learning is one of the most effective in-context learning methods to guide LLM to learn the patterns from a few demonstration examples and to generate similar outcomes like the examples. Here, it is critical to provide examples that are relevant to the questions of interest [1]. However, in our application on AD combinatorial therapy discovery, the therapeutic and their associated biological mechanisms are very diverse; thus, randomly selected examples may fall short of providing relevant information for LLM to learn the patterns to generate response. For example, the question *Q* from (*‘Galantamine*,’ *‘Nicotine*,’ ‘*ICR mice*’) is more similar to one from (*‘Galantamine,’ Memantine,’ ‘ICR mice*’) than one from (*‘Scyllo-inositol,’ ‘neotrofin,’ ‘TgCRND8’*).

Thus we select the most similar question *q* in the learning examples and its associated reasoning *Cq*, and leverage them for inductive reasoning in the inference phase. We derive textual embedding *C_q_* of the question *Q* of interest and *Cq* of the question *Q* in the learning examples using OpenAI’s text-embedding-ada-002 [1]. We then calculated cosine similarity < *Cq*, *C*_*q*_ >/(||*Cq*|| ⋅ ||*C*_*q*_||) to identify the top five similar questions *Q* with the highest similarity along with its correctly generated hypothesis (*Cq*, *Aq*). We appended similar learning examples to the prompt for *Researcher*. So the input for the *Researcher* consists of similar learning examples, question *Q*, and external biomedical knowledge *B*_*Q*_. Note that we guided LLM to have a series of CoT reasoning not only by simply encouraging LLM to “*think step by step,*” but by providing the exact CoT demonstration *C*_*Q*_ in the learning example in the few-shot learning. See the full prompt and the response in Supplementary G.

#### Self-consistency via ensemble

To increase the reliability of LLM’s prediction, we generate the response (*C*_*Q*_, *A*_*Q*_) multiple times. We aggregate them by obtaining consensus prediction *A*^∗^_*Q*_ via majority vote and selecting the most detailed (thus longest) chain of thought *C*^∗^_*Q*_if its paired answer *Aq*is the same as the majority. This ensemble technique can minimize the risk of incorrect prediction by cross-verifying multiple outputs [21].

### Revision phase

#### Evaluate

The theoretical inductive reasoning inevitably carries uncertainty. It is critical to independently evaluate the validity of hypotheses and revise accordingly. After we obtain the hypothesis on efficacy *A*^∗^_*Q*_ and reasoning *C*^∗^_*Q*_ in the inference phase, reviewers need to critically evaluate whether the hypothesis is logical and reasonable. This review process should be independent, thus we used another LLM with comparable performance to *Researcher* (GPT-4), Claude-3-opus, to enhance the independence of the reviewing process.

#### Tree-of-thoughts (ToT)

Reviewers should have diverse perspectives than researchers to critically evaluate the researcher’s hypothesis and identify potential pitfalls that the researchers could not spot. Thus we encourage *Reviewer* to have multiple perspectives and discuss different branches of thoughts via tree-of-thoughts reasoning. ToT reasoning is to generate multiple potential branches of reasoning for a given problem [20]. This approach explores different possibilities and then converges on the most optimal solution. We prompted the *Reviewer* by instructing, “*Imagine three different experts who are in therapy development for Alzheimer’s disease, are tasked with critically reviewing the reasoning…*” (Supplementary H) and provide specific tasks by prompting as “*Please evaluate the response. Explore the potential for drug interactions that could limit or enhance effectiveness*.”

#### Revise

Once reviewers finish the discussion and provide feedback *F*_*Q*_, the moderator aggregates the reviewer’s feedback and researcher’s hypothesis to obtain the final decision. *Moderator* takes input of *Q*, *C*^*^_*Q*_, *A*^*^_*Q*_*F*_*Q*_and deduce the final revised reasoning *C*^*^_*Q*_and answer *A*^*^_*Q*_. See the full prompt and the response in Supplementary I.

##### Algorithm 1 Framework for Drug Combination Identification in Alzheimer’s Disease

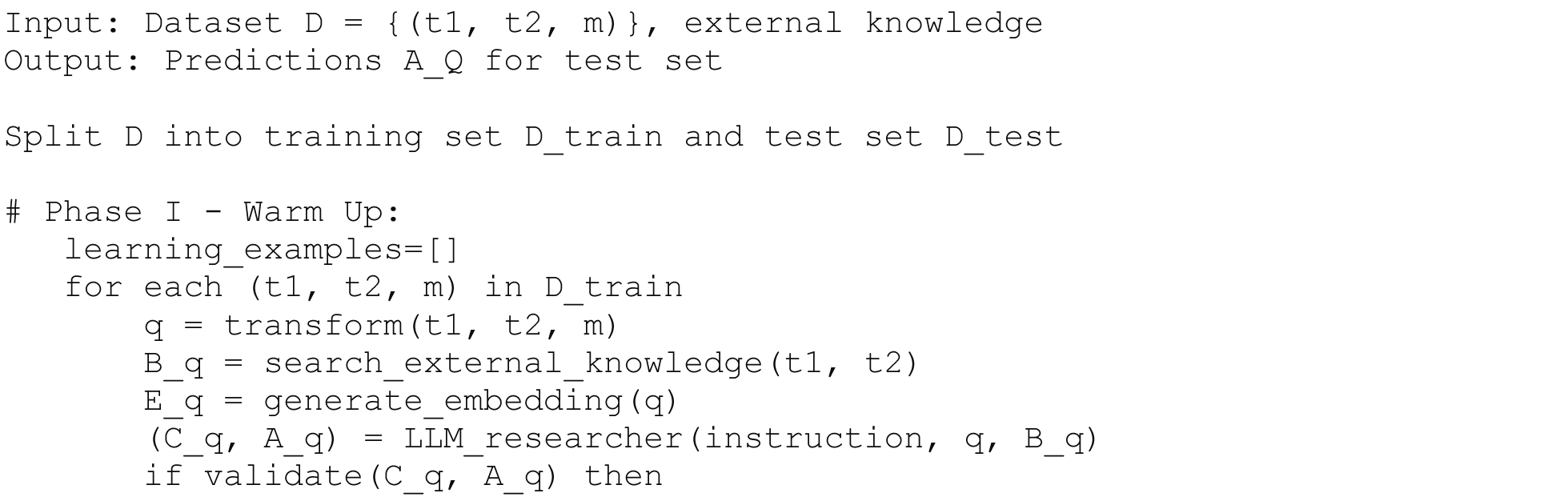
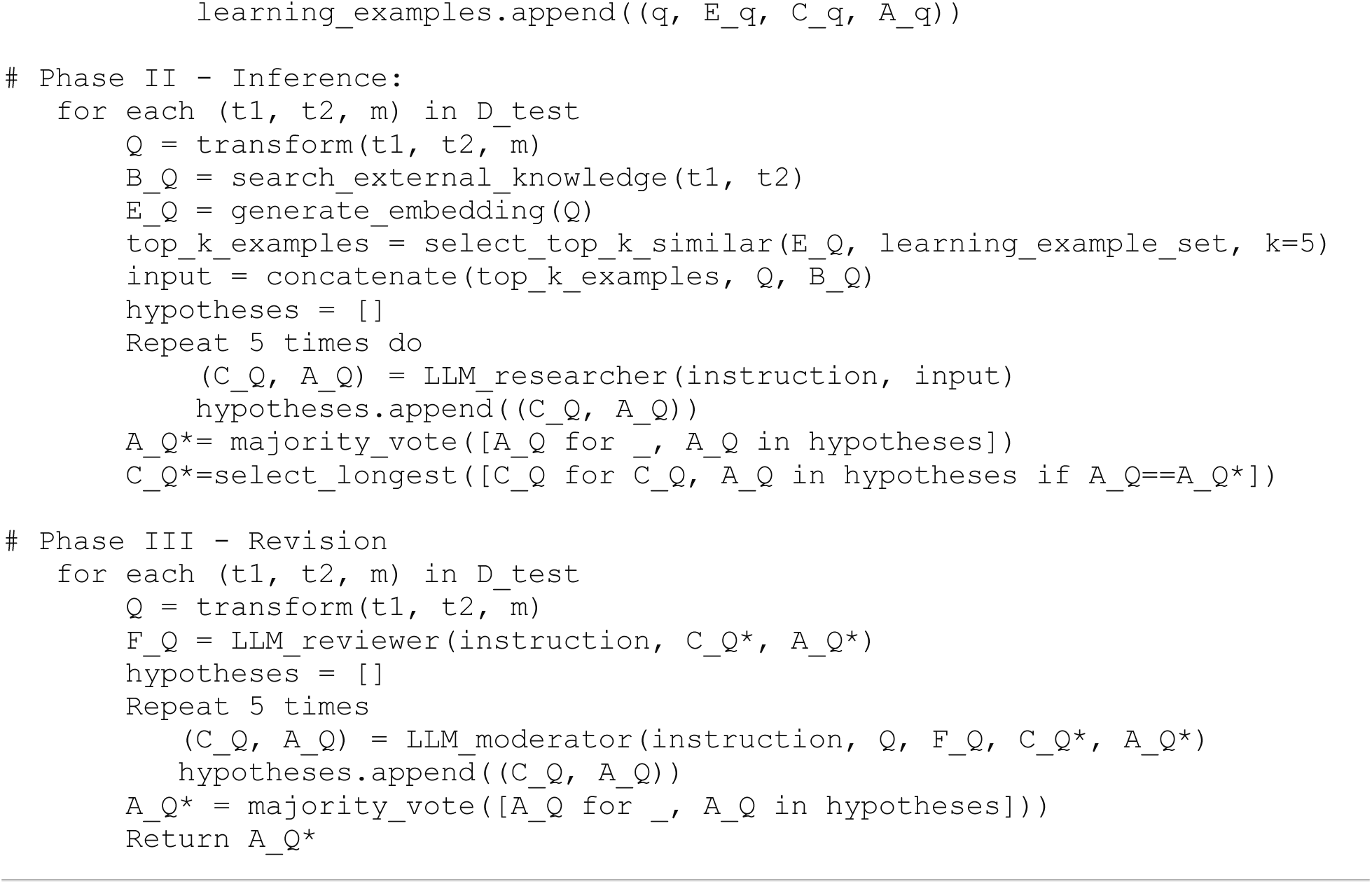

#### Baselines

Due to the lack of sufficient data, data-driven models are not appropriate. Instead, we developed a rule-based baseline model to predict the efficacy of drug combinations. We utilize complementary exposure patterns [36], stating that drug combination is therapeutically effective if the targets of the drugs hit the disease module without overlap (Algorithm 2). The target genes of the drugs were collated from multiple sources, including Drug Target Commons, PubChem, and CTDbase [23,37,38], whereas AD-related genes were derived from Agora’s nominated gene list [39]. Within the test set, target gene information was unobtainable for 76 drugs (34.3%). Additionally, 76 drug combinations (48.72%), can not be evaluated using the baseline model due to the absence of necessary target gene information.

##### Algorithm 2 Framework for Baseline Model

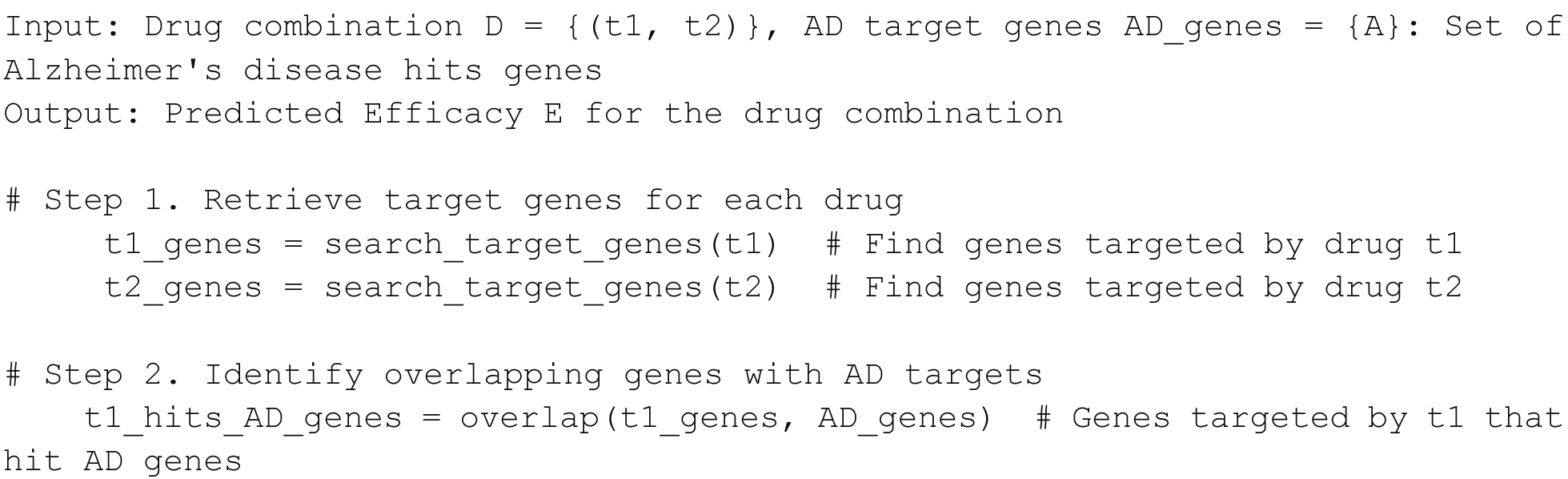
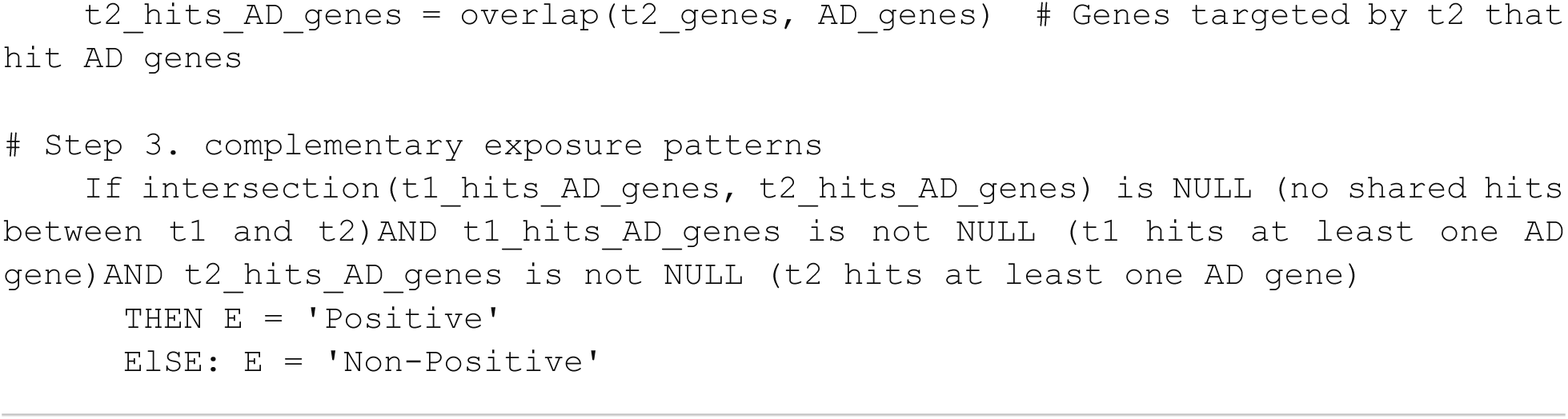

### Supplementary Section

#### A. Problem formulation

Our objective is to predict whether a combination of therapeutic agents *t*_1_ and *t*_2_ have a positive efficacy *y* when tested in an in vivo model *m*. That is, we aim to develop a model *f* such that *y* = *f*(*x*), where *x* is a triplet of (*t*_1_, *t*_2_, *m*). Here, *t*_1_, *t*_2_, and *m* are not only drawn from a finite set, but can be a new or investigational therapeutic agent (e.g. Membrane-free *stem cell extract*) or in vivo model (e.g. “*Rats induced with AD using aluminum chloride*,” “*Mice induced with cerebral malaria”*), which are not registered with a formal identifier, thus best described as a natural text. We convert the structured input (*t*_1_, *t*_2_, *m*) into a natural text question *Q*, following [9]. For example, we convert the combination (‘Galantamine,’ ‘Nicotine,’ ‘ICR mice’) into ‘*Decide if the combination of Galantamine and Nicotine is effective or not to treat ICR mice model in theory.’*

In some previous studies [9], the effectiveness of combinations of therapeutic agents was measured as synergy. However, the synergy requires dose-dependent inhibition, which is not available in an in vivo model. As most in vivo experiments only report efficacy (without formal calculation of synergistic effect), our focus is also on efficacy (rather than synergy). Rather than a specific efficacy measurement, we focused on a broad sentiment as an efficacy measurement (positive efficacy or not) because this is more transferable to different studies.

#### B. Data collection

We collect scientific articles that report the efficacy of therapeutic agent combinations on AD in vivo models. We first utilized the Alzheimer’s Disease Preclinical Efficacy Database [40], a data resource dedicated to the preclinical efficacy studies of candidate therapeutics for Alzheimer’s Disease. Among the 1,463 articles in AlzPED, we manually reviewed and selected 39 articles that experimented on more than two therapeutic agents.

We further searched for more related articles based on the 39 selected articles. We crafted a series of PubMed search queries for AD *and* one of its mechanisms (e.g., ‘amyloid beta,’ ‘tau,’ ‘inflammation,’ ‘cognitive dysfunction,’ ‘oxidative stress’), *and* combination (e.g., ‘co-administration’), *and* in vivo models (e.g., ‘mouse,’ ‘rat’). Specific search queries are available in Supplementary E. We extracted 376 additional articles meeting the query from PubMed. 10 out of 39 (25.642) AlzPED articles were searchable from the query.

We then extracted therapeutic agents, in vivo models, and their efficacy from the abstract of the selected articles. We excluded articles in which drugs were used to induce AD or suppress mechanisms for mechanistic study. Among the 376 articles, 206 articles reported positive efficacy, and 3 reported mixed or partial effects, 16 reported negative efficacy. All others are not relevant.

#### C. Data augmentation

There was an imbalance in the number of samples reporting positive and negative efficacy. Researchers, of course, tend to publish positive efficacy more, thus the collected data is inevitably biased toward combinations reporting positive efficacy. In addition to combinations reporting non-positive efficacy in the literature, we create plausible samples with unknown efficacy (unlabeled data). We use these unlabeled samples as non-positive samples with noise and use them in both the Warm-up phase and Inference phase. The non-positive samples were created by randomly replacing either one of the drugs or an in vivo model from the positive efficacy combinations. For example, given an efficacious combination *(Acamprosate, Baclofen, mThy1-hAPP751 (TASD41))*, we created a non-positive combination by replacing Baclofen. As a result, we have *(Acamprosate, Melatonin, mThy1-hAPP751 (TASD41))*.

#### D. Evaluation

We evaluate whether the prediction of Coated-LLM is accurate by comparing the binary prediction (i.e., positive v.s. non-positive) with the ground-truth label. We reported accuracy rate, precision, and recall. We first evaluated the accuracy via cross-validation using the test set and via external validation using in-house private data. The external dataset has 11 drug combinations, of which 9 is labeled as non-positive efficacy. Furthermore, during the evaluation of the external data, we augmented the initial learning examples from the Warm-up phase by incorporating combinations that were correctly predicted during the Revision phase. After augmenting the learning examples, we had 346 combinations (195 combinations with positive efficacy; 151 showing non-positive efficacy) serving as learning examples for predicting efficacy on our private data.

We conducted an ablation study to understand the relative contributions of each component in our model. We iteratively introduced each component and measured the performance differences. Since these components are not statistically independent [1,2], we should consider the performance differences as the components’ relative contributions.

#### E. PubMed search query to extract relevant scientific literature

(“Alzheimer”) AND (“Aβ” OR Abeta OR “Amyloid-β peptide” OR “Beta-amyloid” OR “Amyloid” OR “Aβ” OR “Amyloidogenic” OR “Cerebral amyloidosis” OR “antioxidant” OR “tau” OR “inflammation” OR “cognitive” OR “synaptic”) AND (“combination therapy” OR “combination treatment” OR “co-administration” OR “co-treatment” OR “dual therapy” OR “multi-therapy” OR “adjunct therapy” OR “combined treatment” OR “therapy plus”) AND (mice OR mouse OR rat OR “Morris water maze” OR animal) NOT (review[Publication Type] OR “review” OR “perspective” OR “clinical trial” OR “Randomized controlled trial”)’

#### F. Prompt and generated response in the Warmup phase

**System:** You are an expert in therapy development for Alzheimer’s disease and you are trying to decide if the combination of two drugs is effective or not to treat or slow the progression of Alzheimer’s disease in theory. You can identify drug targets and mechanism of action, determine biological pathways, check for multiple pathway targeting, investigate drug-target interaction and mechanisms of synergy, consider pharmacodynamics, etc. Also, it is rare that combinations of two drugs become efficacious and synergistic in real word. As a proficient neurobiologist, use your own knowledge and search for external information if necessary.

**User:** Background: <Background> {Pathway Information}</Background>. Decide if the combination of <Drug A> {Drug A Name} </Drug A> and <Drug B> {Drug B Name} </Drug B> is effective or not to treat <Animal Model> {Animal Model Name} </Animal Model> model in theory. Take a breath and work on this problem step by step. And conclude using the format: “Effective in theory: <Positive or Non-positive>”.

List 2.1. Prompt for *Researcher* LLM in the Warm-up phase.

First, let’s understand the individual drugs and their action mechanisms.

(-)-Epigallocatechin-3-gallate (EGCG) is the main polyphenolic compound present in green tea, and it has been shown to have various health benefits, including neuroprotective effects. EGCG aids in reducing beta-amyloid production and aggregation, a key pathological marker of Alzheimer’s disease. It does this by modulating the activity of gamma-secretase, an enzyme involved in beta-amyloid production. It also has anti-inflammatory and antioxidant properties which can protect neurons from damage.

Ferulic Acid (FA), on the other hand, is a phenolic compound found in the cell walls of plants. It has antioxidant properties and has been shown to protect against beta-amyloid-induced neurotoxicity. FA has been found to inhibit tau protein hyperphosphorylation, another hallmark of Alzheimer’s disease. It also has anti-inflammatory effects.

Looking at the APPswe/PSEN1dE9 mouse model of Alzheimer’s, it overproduces beta-amyloid due to mutations in the amyloid precursor protein (APP) and presenilin 1 (PSEN1) genes. Therefore, a treatment that can reduce beta-amyloid production and aggregation as well as tau hyperphosphorylation could potentially be beneficial.

Combining EGCG and FA could potentially target multiple pathways involved in Alzheimer’s pathogenesis. EGCG could reduce beta-amyloid production and aggregation, while FA could inhibit tau hyperphosphorylation. Both have anti-inflammatory and antioxidant effects that could protect neurons from damage. This multiple pathway targeting could potentially lead to a synergistic effect, although this would need to be confirmed experimentally, as drug interactions can be complex and unpredictable.

In terms of pharmacodynamics, both EGCG and FA are well-absorbed and can cross the blood-brain barrier, which is crucial for any drug designed to treat a neurological disorder like Alzheimer’s.

Therefore, in theory, the combination of EGCG and FA could potentially be effective in treating the APPswe/PSEN1dE9 mouse model of Alzheimer’s disease. However, it’s important to note that this is theoretical, and further research, including pre-clinical and clinical trials, would be necessary to confirm this.

Effective in theory: Positive.

List 2.2. Example of generated response from *Researcher* LLM in the Warm-up phase.

#### G. Prompt and generated response in the Inference phase

**System**: You are an expert in therapy development for Alzheimer’s disease and you are trying to decide if the combination of two drugs is effective or not to treat or slow the progression of Alzheimer’s disease in theory. Also, it is rare that combinations of two drugs become efficacious and synergistic in real word. As a proficient neurobiologist, use your own knowledge and search for external information if necessary.

**User**: <Question 1> {Question 1} </Question> : <CoT 1> {Reasoning 1} </CoT 1>

<Question 2> {/Question 2} <2> : <CoT 2> {Reasoning 2} </CoT 2>

<Question 3> {/Question 3} <3> : <CoT 3> {Reasoning 3} </CoT 3>

<Question 4> {/Question 4} <4> : <CoT 4> {Reasoning 4} </CoT 4>

<Question 5> {/Question 5} <5> : <CoT 5> {Reasoning 5} </CoT 5>

<Background> {Pathway Information} </Background>

<Test Question> {Test Question} </Test Question>

Take a breath and work on this problem step by step. And conclude using the format ’Effective in theory: <Positive or Non-positive>.’

List 3.1. Prompt for *Researcher* LLM in the Inference phase.

Reasons: Galantamine is already known for its use in Alzheimer’s disease treatment. It works by inhibiting acetylcholinesterase, an enzyme that breaks down acetylcholine in the brain. By inhibiting this enzyme, Galantamine increases the concentration of acetylcholine, which can improve cognitive function.

Mecamylamine is a nonselective and noncompetitive antagonist of the nicotinic acetylcholine receptors that has been used as a ganglionic blocker in hypertension. However, in the context of Alzheimer’s disease, there is not much evidence to support its use. Its mechanism of blocking nicotinic acetylcholine receptors could counteract the positive effects of Galantamine, which is aiming to increase acetylcholine activity.

In the ICR (beta Amyloid 25-35 Peptide Injection Model) mouse model, the combination of these two drugs might not be effective. Since Mecamylamine is a nonselective antagonist, it could block the effects of increased acetylcholine caused by Galantamine. This could lead to a reduction in the overall effectiveness of the therapy.

In terms of pharmacodynamics, the two drugs could potentially interact in a way that would reduce their overall effectiveness. Additionally, the side-effect profiles of these two drugs could potentially overlap, leading to an increase in adverse effects.

Considering the above information, the combination of Galantamine and Mecamylamine is likely not effective in treating or slowing the progression of Alzheimer’s disease, especially in an ICR (beta Amyloid 25-35 Peptide Injection Model) mouse model.

Effective in theory: Non-positive.

List 3.2. Example of generated response from *Researcher* LLM in the inference phase.

#### H. Prompt and generated response in the Revision (evaluate) phase

**System**: Imagine three different experts who are in therapy development for Alzheimer’s disease, are tasked with critically reviewing the reasoning and conclusions regarding the effectiveness of a combination of two drugs on an Alzheimer’s disease animal model from a theoretical perspective. All experts will write down 1 step of their thinking, then share it with the group. Then all experts will go on to the next step, etc. If any expert realizes they’re wrong at any point then they leave. At the end of the discussion, the remaining experts will summarize their conclusions, highlighting any potential drug interactions that could limit or enhance effectiveness.

**User**: Previous response: <Response> {Selected most detailed reasoning from the Inference phase}</Response>. Please evaluate the response. Explore the potential for drug interactions that could limit or enhance effectiveness.

List 4.1. Prompt for *Reviewer* LLM in the Revision (evaluate) phase.

Expert 1: The reasoning about the mechanism of action of Galantamine and Mecamylamine is correct. Galantamine increases acetylcholine levels by inhibiting its breakdown, while Mecamylamine blocks nicotinic acetylcholine receptors. This could lead to counteracting effects.

Expert 2: I agree with Expert 1. Additionally, we should consider the potential for pharmacokinetic interactions. Both drugs are metabolized by the liver, which could lead to changes in their metabolism and elimination when administered together. This could affect their effectiveness or increase the risk of side effects.

Expert 3: The point about Mecamylamine’s anticholinergic side effects potentially exacerbating symptoms in Alzheimer’s patients is valid. However, we should also consider the potential for beneficial interactions. Some studies have suggested that nicotinic acetylcholine receptor antagonists like Mecamylamine could have neuroprotective effects by reducing inflammation and oxidative stress in the brain.

Expert 2: While that’s an interesting point, the evidence for neuroprotective effects of Mecamylamine in Alzheimer’s disease is limited. The potential for it to counteract the benefits of Galantamine by blocking acetylcholine receptors likely outweighs any potential neuroprotective effects.

Expert 1: I agree. The conclusion that the combination of Galantamine and Mecamylamine is likely not effective in treating Alzheimer’s disease in the ICR mouse model seems reasonable based on the available evidence.

Expert 3: I concur with the conclusion. The potential for Mecamylamine to negate the benefits of Galantamine by blocking acetylcholine receptors, along with the increased risk of side effects due to pharmacokinetic interactions, suggests that this combination is not likely to be effective. Summary: Based on the mechanism of action of Galantamine and Mecamylamine, their potential to counteract each other’s effects, and the increased risk of side effects due to pharmacokinetic interactions, the combination of these drugs is likely not effective in treating Alzheimer’s disease in the ICR mouse model. While there is some evidence for potential neuroprotective effects of nicotinic acetylcholine receptor antagonists like Mecamylamine, this is outweighed by the potential for it to negate the benefits of Galantamine by blocking acetylcholine receptors.

List 4.2. Example of generated response from *Reviewer* LLM in the Revision (evaluate) phase.

#### I. Prompt and generated response in the Revision (revise) phase

**System**: You are an expert in therapy development for Alzheimer’s disease and you are trying to decide if the combination of two drugs is effective or not to treat or slow the progression of Alzheimer’s disease in theory. Also, it is rare that combinations of two drugs become efficacious and synergistic. As a proficient neurobiologist, use your own knowledge and search for external information if necessary.

**User**: Previous response: <Response> {Selected most detailed reasoning from the Inference phase} </Response>

Feedback: <Feedback> {Detailed Feedback} </Feedback>

Based on the previous response and feedback, <Question> {Test Question} <>. Take a breath and work on this problem step by step. And conclude using the format ’Effective in theory: <Positive or Non-positive>.’

List 5.1. Prompt for *Moderator* LLM in the Revision (revise) phase

Okay, let’s break this down step by step:

1. Galantamine is an acetylcholinesterase inhibitor, which increases acetylcholine levels in the brain by preventing its breakdown. This can potentially improve cognitive function in Alzheimer’s patients.
2. Mecamylamine is a non-selective, non-competitive antagonist of nicotinic acetylcholine receptors. It blocks the action of acetylcholine on these receptors.
3. When used together, Mecamylamine could potentially counteract the beneficial effects of Galantamine by blocking the acetylcholine receptors that Galantamine is trying to stimulate by increasing acetylcholine levels.
4. Mecamylamine also has anticholinergic side effects, which could exacerbate symptoms in Alzheimer’s patients.
5. Both drugs are metabolized by the liver, which could lead to pharmacokinetic interactions and affect their effectiveness or increase the risk of side effects when used together.
6. While there is some evidence for potential neuroprotective effects of nicotinic acetylcholine receptor antagonists like Mecamylamine, this is likely outweighed by its potential to negate the benefits of Galantamine.
7. The ICR (beta Amyloid 25-35 Peptide Injection Model) mouse model is used to study Alzheimer’s disease. The effectiveness of the combination therapy in this model would depend on how well it addresses the pathological changes induced by the beta-amyloid peptide injection.
8. Given that Mecamylamine could counteract the effects of Galantamine, increase the risk of side effects, and potentially exacerbate Alzheimer’s symptoms, it is unlikely that the combination would be effective in treating the ICR mouse model of Alzheimer’s disease.

Therefore, based on the available evidence and considering the potential interactions between Galantamine and Mecamylamine, their mechanisms of action, and the pathology of the ICR mouse model, the combination of these drugs is likely not effective in treating Alzheimer’s disease in this model.

Effective in theory: Non-positive.

List 5.2. Example of generated response from *Moderator* LLM in the Revision (revise) phase.

#### J. Ablation study only using data from literature mining

The ablation study which we only used data from literature mining in both the Warm-up and the Inference and Revision phases illustrated similar trends.

**Figure S1.**
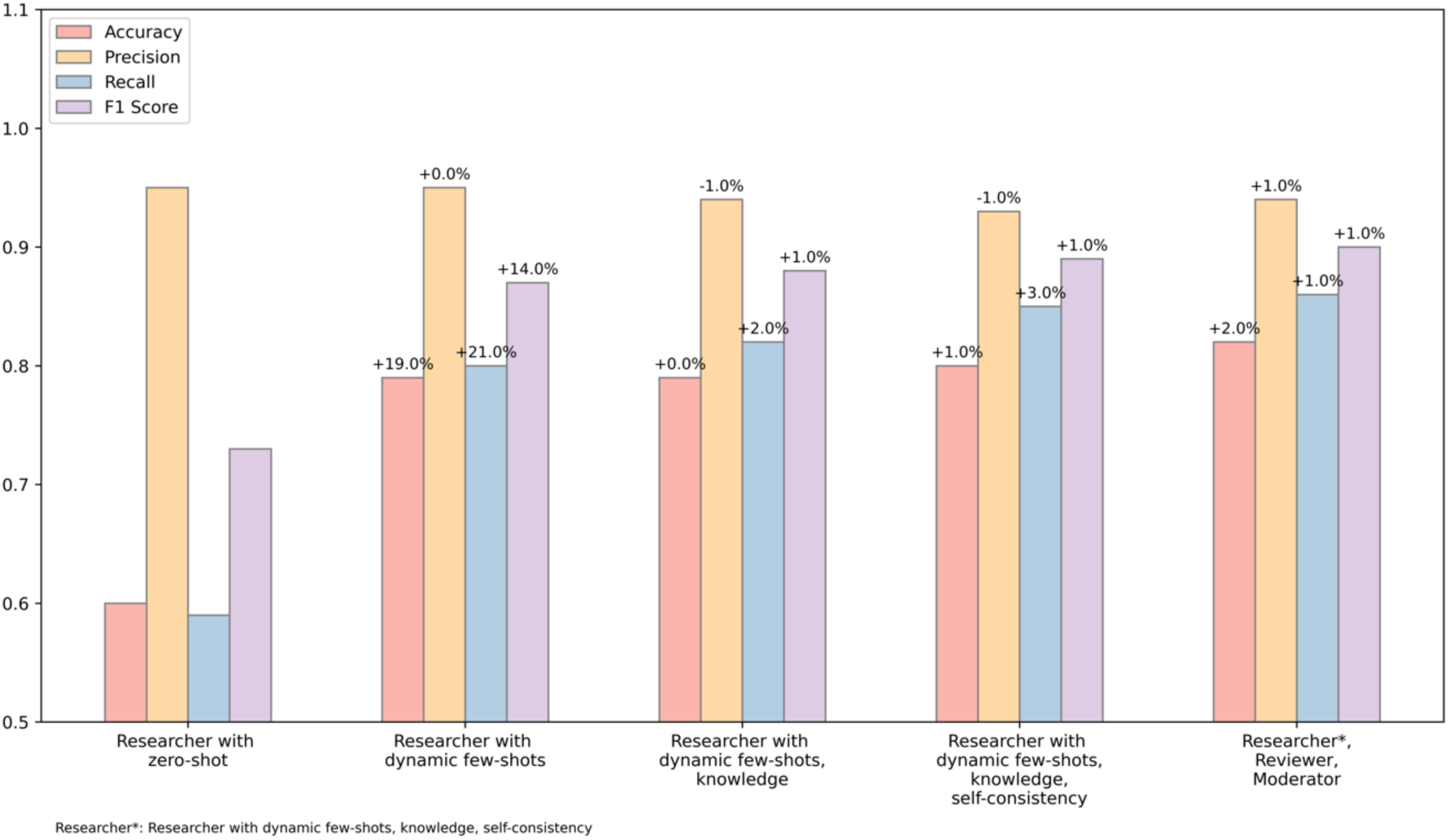
Visual illustration of Coated-LLM components and additive contributions to the performance (only use literature mining data). Coated-LLM combines kNN-based five-shots dynamic learning example selection, external pathway knowledge, self-consistency (n=5), *Reviewer*, and *Moderator*.

**Table S1.** Contingency table of prediction outcomes with examples using the baseline model.

| **Drug A** | **Drug B** | **Animal model** | **Predicted efficacy** | **Actual efficacy (reference)** |
|---|---|---|---|---|
| **True positive (n=6)** |  |  |  |  |
| Pyrroloquinoline Quinone | NT-020 | 5xFAD | Positive | Positive [41] |
| Donepezil | Pentoxifylline | CuSO4 intake in experimental rats | Positive | Positive [42] |
| **False positive (n=7)** |  |  |  |  |
| Levetiracetam | Donepezil | Aged C57/Bl6J | Positive | Non-positive (Augmented data) |
| Boron | Ferulic Acid | Wistar Albino Male Rats injected stereotaxically with STZ | Positive | Non-positive (Augmented data) |
| **False negative (n=31)** |  |  |  |  |
| CCMI | Donepezil | scopolamine-treated rats | Non-positive | Positive [43] |
| Rosiglitazone | Vorinostat | Intracerebroventricular streptozotocin induced model of AD in mice | Non-positive | Positive [44] |
| **True negative (n=36)** |  |  |  |  |
| Cholesterol | Homocysteine | Sprague Dawley rats | Non-positive | Non-positive [29] |
| Dexamethasone | Tofacitinib | 5xFAD | Non-positive | Non-positive [45] |

**Table S2.** Contingency table of prediction outcomes with examples using the baseline model for the external data.

| **Drug A** | **Drug B** | **Model** | **Predicted efficacy** | **Actual efficacy** |
|---|---|---|---|---|
| **True positive (n=0)** |  |  |  |  |
| **False positive (n=6)** |  |  |  |  |
| Rivastigmine | Vitamin E | HT22 Mouse Hippocampal Neuronal Cell Line | Positive | Non-positive |
| Memantine | Tolcapone | HT22 Mouse Hippocampal Neuronal Cell Line | Positive | Non-positive |
| **False negative (n=2)** |  |  |  |  |
| Galantamine | Mifepristone | HT22 Mouse Hippocampal Neuronal Cell Line | Non-positive | Positive |
| Galantamine | Caffeine | HT22 Mouse Hippocampal Neuronal Cell Line | Non-positive | Positive |
| **True negative (n=3)** |  |  |  |  |
| Galantamine | Diclofenac | HT22 Mouse Hippocampal Neuronal Cell Line | Non-positive | Non-positive |
| Donepezil | Octyl-methoxycinnamate | HT22 Mouse Hippocampal Neuronal Cell Line | Non-positive | Non-positive |

## Data Availability

The data generated from literature mining and drug hit AD genes (baseline model) for this study can be accessed via the following link: AD Efficacy Combinatorial Therapy

## Code Availability

The code for Researcher, Reviewer, and Moderator can be found at https://github.com/QidiXu96/Coated-LLM

